# Improving capacity for advanced training in obstetric surgery: Evaluation of a blended learning approach

**DOI:** 10.1101/2023.04.25.23289116

**Authors:** Helen Allott, Alan Smith, Sarah White, Irene Nyaoke, Ogoti Evans, Michael Oriwo Oduor, Steven Karangau, Sheila Sawe, Nassir Shaaban, Ochola Ephraim, Charles Anawo Ameh

**Affiliations:** Emergency Obstetric Care and Quality of Care Unit, Liverpool School of Tropical Medicine (LSTM), Liverpool, United Kingdom; Liverpool School of Tropical Medicine Kenya, Nairobi, Kenya; Directorate of Reproductive Health, Moi University Teaching and Referral Hospital (MTRH), Eldoret, Kenya; Bondo Subcounty Hospital, Bondo, Kenya; Muriranjas Subcounty Hospital, Murang’a, Kenya; University of Nairobi, Nairobi, Kenya

## Abstract

**Introduction:** Significant differences in outcomes for mothers and babies following obstetric surgical interventions between low- and middle-income countries and high-income settings have demonstrated a need for improvements in quality of care and training of obstetric surgical and anaesthetic providers. To address this a five-day face-to-face training intervention was developed. When the COVID-19 pandemic interrupted its roll-out, the course was redesigned for delivery by blended learning.

**Methods:** This 3-part blended-learning course (part-1: 15 hours self-directed online learning, part-2: 13 hours facilitated virtual workshops and part-3: 10 hours face-to-face delivery), was conducted in Kenya. We assessed the completion rate of part-1 (21 assignments), participation rate in parts 2 and 3, participant satisfaction, change in knowledge and skills and compared the cost of the blended delivery compared to the 5-day face-to-face delivery, in GB Pounds.

**Results:** 65 doctors took part in part 1, 53 completing at least 90% of the assignments. 60 doctors participated in part 2, and 53 participated in part 3. Participants completing an evaluation reported (n=53) attending the training was a good use of their time (each of parts-1 and 3: 98%, part-2: 94%) and would recommend this to other colleagues (part-1 and 3: 98%, part-2: 90%). Mean (SD) knowledge score improved from 64% (7%) to 80% (8%) and practical skills from 44% (14%) to 87% (7%). The blended course achieved a cost-saving of £207 per participant compared to the 5-day face-to-face delivery approach.

**Conclusion:** We have demonstrated that a blended learning approach to clinical training in a low resource setting is feasible, acceptable and more cost effective. More studies are required to investigate the effectiveness of this approach on health outcomes.

## Introduction

Improving access to quality maternal and newborn health services, while addressing context specific challenges, is critical to meeting the global maternal health Sustainable Development Goal target of less than 70 maternal deaths per 100,000 live births by 2030 (1). Caesarean section (CS) is a life-saving emergency obstetric care major surgical procedure that, if performed when indicated, should reduce the risk of maternal and perinatal mortality. Boerma and colleagues reported in 2018 that 21.1% (29.7 million) of births that occurred globally in 2015 were by CS. There was also an increasing trend in CS rates in some world regions and in others it was still suboptimal (2). Population based CS rates above 20% have not been shown to improve perinatal or neonatal outcomes (3) and rates of less than 10%, as seen in many low-income and middle-income countries, indicate inadequate access to medically indicated CS (4). Short and long-term risks to CS have been reported, the CS related maternal mortality ratio in LMICs is 100 times more than in high income countries and a high stillbirth rate associated with CS of 82.5 per 1, 000 has been reported (5–9).

Interventions to reduce unnecessary CS need to address the drivers of excessive and inappropriate use of CS at three levels: a) childbearing women, families, communities and broader society, b) health professionals c) health systems, financial reimbursements, organisational design and cultures. These interventions include clinical and non-clinical interventions to achieve meaningful reductions in the risk from CS (10). The systematic review by Sobhy and colleagues identified 3 themes to be addressed for optimal outcomes from caesarean sections: 1) access to safe surgery, 2) the management of perioperative complications in Low and Middle Income Countries (LMICs) and 3) labour related complications related to the second stage of labour. Caesarean sections performed in the second stage of labour can be very challenging especially for less experienced doctors. Assisted vaginal birth with vacuum delivery device, when indicated, may be a safer option. In a multi-country review of assisted vaginal delivery, Bailey et al found that one of the prime reasons for low levels of assisted vaginal delivery in LMICs was a lack of staff training, underlining the need to provide enhanced training in this procedure (11).

The first Confidential Inquiry into Maternal Deaths in Kenya report, published in 2018, showed that 37% of women who died had had a caesarean section. Delays in starting treatment, inadequate clinical skills and inadequate monitoring were found to be the significant health workforce factors associated with these deaths (9). Therefore, in 2018 we designed a health worker training intervention that addressed these factors, plus the key themes identified in a systematic review (7) and the lack of skills to perform assisted vaginal birth. When the covid-19 pandemic rendered it impossible to continue with delivering this training face-to-face, we adapted the course delivery to suit a blended learning delivery model. Blended learning has been described as “learning that happens in an instructional context which is characterized by a deliberate combination of online and classroom-based interventions to instigate and support learning” (12). Successful blended learning requires the integration of both virtual and face-to-face methods, rather than merely using the on-line component as an add-on to classroom teaching.

The objective of this paper is to describe and evaluate the blended learning version of Liverpool School of Tropical Medicine’s Advanced Obstetric Surgical and Anaesthetic Care training package.

## Methods

### Study setting

We recruited 69 potential participants from Ministry of Health public hospitals in the five counties in Kenya (Garrisa, Taita Taveta, Kilifi, Usain Gishu and Vihiga) supported by the United Kingdom’s Foreign and Commonwealth Development Office Kenya’s Maternal and Newborn Health (MNH) programme. All participants were medical doctors with experience ranging from first year interns (four) through to consultants (eleven) and residents in training (four), but the majority were medical officers (MOs), four of whom had six or more years-experience. Forty-six MOs had been in post for between one and five years. In sub-county and county hospitals medical officers are often responsible for providing obstetric care without consultant supervision.

### Intervention

The LSTM Advanced Obstetric Surgical and Anaesthetic Course (AOAC) was created based on our experience designing, implementing and evaluating the LSTM Emergency Obstetric and Newborn Care training package (13-17). Both courses are based on principles of adult learning and a structured approach to teaching (18).

The objectives of the AOAC are to improve capacity 1) for decision making, and leadership of maternity care teams, 2) to provide quality perinatal care, 3) to perform caesarean sections safely, and 4) to perform assisted vaginal birth, by addressing the topics listed in Box 1. Facilitator and participant manuals are made available to facilitate learning and standardise teaching (19, 20).

#### Box 1

Key topics covered in the LSTM AOAC course.

- Improving decisions for operative obstetric births
- Labour ward leadership and management
- Improving interdisciplinary communication skills
- Safe anaesthesia and surgery
- Enhanced surgical skills for routine and complex caesarean surgery.
- Advanced skills in assisted vaginal delivery and perineal trauma repair.
- Detection and management of post-operative complications including use of obstetric early warning system.
- Patient counselling skills, consent
- Improving quality of care, respectful maternity care
- Performing clinical audit, maternal death surveillance and response
- Action planning

This course, originally designed and delivered as an entirely face to face intervention for up to 28 participants, followed a journey. It commenced with decision-making for operative birth, then patient consent and preparation for surgery and safe surgical and anaesthetic practices for Caesarean section. There was also an emphasis on assisted vaginal birth techniques and post-operative care, management of complications, patient counselling for future birth and the use of audit as a quality improvement tool. The training culminated with participants working in functional teams to develop implementation plans and strategies to put their preferred changes, based on their learning, into practice upon their return to work.

For the blended learning version, we reduced the face-to-face component to a 1.5-day duration focused on skill acquisition using models. The course lectures were recorded and divided into smaller sections interspersed with on-line activities, to encourage user participation, and clinical scenarios, role-plays and workshops were adapted for on-line discussion groups.

### Standard training package

The standard 5-day AOAC package developed and piloted in Cambodia, Nigeria and Kenya before the COVID 19 pandemic, consisted of brief lectures (31% or 10 hours of the whole course) interspersed with interactive small group activities (39% or 13 hours of the whole course) such as facilitator led interactive workshops, simulation-based learning (clinical scenarios and role playing) and practical skills sessions (30% or 10 hours of the whole course) with the use of obstetric, anaesthetic and newborn care mannequins (Table 1). The objective of the five-day interactive training course, targeted at obstetric and anaesthetic service providers, was to enhance decision making, teamwork and practical skills in obstetric surgery and anaesthesia, to improve maternal and newborn outcomes.

**Table 1:** Comparison of duration of each training approach and trainer requirement.

|  | Duration in hours (% of total) |  |  |
| --- | --- | --- | --- |
|  | Learning approach | Standard 5-day face-to-face approach | Alternative blended learning approach |
| **Knowledge** (face-to-face, self-directed or virtual) | Lectures | 10 (31%) | 15 (40%) |
|  | Workshops | 13 (39%) | 12.5 (33%) |
| **Practical skills** (face-to-face) | Simulation Based Education, hands-on training | 10 (30%) | 10 (27%) |
| **Total duration in hours (days)** |  | 33 hours (5 days) | 37.5 (5 days) |
| **Facilitator/participant ratio for practical sessions** |  | 1:4 | 1:4 |
| **Total number of facilitators per practical session** |  | 8 | 6 |

The face-to-face course was attended by a multidisciplinary group of skilled health workers including obstetricians, medical officers and obstetric trainees, anaesthetists and peri-operative care nurses. The obstetricians, medical officers and obstetric trainees attended for the full five days and anaesthetists/peri-operative nurses for three days. For two days participants from both specialities worked together and on one day they were provided with advanced skills training relevant to their respective specialities.

The training culminated with participants working together in facility teams to formulate action plans based on the training to improve their practice and quality of care provided afterwards.

### Blended training approach

During the Covid pandemic, training was provided purely to obstetric cadres because the anaesthetic trainers were not available due to Covid-related clinical commitments. The blended learning approach consisted of three basic components: self-directed learning consisting of recorded lectures (40% or 15 hours of the whole course), workshops delivered through five real-time interactive small group facilitator lead learning discussions (33% or 12.5 hours of the whole course) followed by 1.5 days of face-to-face practical skills training (27% or 10 hours of the whole course). The blended-learning course takes slightly longer to complete (37.5 hours versus 33 for the face-to-face version. This is due to an estimated longer period to complete the self-directed learning component of listening to the recorded lectures and accompanying activities (Table1).

We used the limited version of the Google® Classroom learning platform to pilot the self-directed component. The lectures from the original course were recorded and uploaded to YouTube® and the duration of time each participant spent listening to each lecture was monitored on the learning platform. To enhance concentration and interest, the 21 lectures were divided into short sections, mostly lasting between 5 to 10 minutes, interspersed with a variety of participatory activities, including short videos and questions. To encourage engagement, at the end of each lecture participants were required to complete a short quiz. Once the quiz had been completed and submitted, participants were enabled to access the correct answers for the questions together with brief explanations regarding the rationale underlying these answers.

The interactive small group guided discussions lasted 2.5 hours each. Participants were divided into eight groups, each with two facilitators leading participants through a pre-prepared script to guide the discussion. Topics covered included respectful care, decision-making skills for operative birth, informed consent, prioritisation and triage on the labour ward, use of the World Health Organisation (WHO) Safe Surgical Check List, safe operating procedures and recognition of the deteriorating patient intra- and post-operatively, managing post-operative patients using an obstetric early warning system, prevention, identification and management of intra- and post-operative haemorrhage and other post-operative complications, safe blood transfusion, analgesia, documentation, counselling for future births, audit and Maternal Death Surveillance and Response together with training on action planning.

The surgical skills selected for the face-to-face training component included assisted vaginal birth, obstetric anal sphincter injury repair, insertion of B-Lynch sutures, improved techniques for knot-tying and uterine incision closure, extraction of an impacted fetal head, balloon tamponade after placenta praevia and venous cut-down, together with revision of airway assessment and management skills, and newborn resuscitation.

### Study design and evaluation framework

We used a cross sectional study design and the adapted Kirkpatrick’s training evaluation framework, previously described to evaluate the participants engagement, acceptance, reaction and immediate change in knowledge and skills (Kirkpatrick level 2) (12). Additionally, we compared the cost in Pounds Sterling per participant of the blended learning approach to the cost of the 5-day face-to-face training approach.

To evaluate participant engagement, we monitored the time that participants spent on the self-directed sessions and compared this to the minimum time necessary to view all the recorded lecture sections and accompanying videos. To evaluate the participant acceptance and reaction to the blended learning approach, they were invited to complete an on-line feedback evaluation, using a Likert-scale together with free comments, at the end of the third component of the training.

Learning was evaluated by means of an overall multiple choice (MCQ) knowledge assessment taken before and after all the course elements. All participants who attended the face-to-face component took part in three objective structured clinical examinations (OSCEs) before and after that part of the training.

### Data collection procedure

Participants were provided with funds for airtime to enable them to engage on the internet-dependent components of the course. They were provided with both written and video instructions explaining how to engage with the self-directed learning components of the course. A dedicated on-line education consultant provided support to any participant who found engagement with the online learning platform problematic.

Authors were aware of participants’ identities because they were teaching them and also conducting face-to-face objective structured clinical examinations as a part of the evaluation. Participants were issued with an ID number and no names were recorded on evaluation score sheets, only ID numbers. Date concerning results of the evaluations was entered onto a password protected file, accessible only to the educational consultant (AS) who was aware of the ID number of each participant. This was used to provide essential feedback to each participant regarding their performance as and when they requested it.

For the on-line group discussions 18 volunteer faculty members were recruited in total, including six from Kenya and 12 from UK. Wherever possible, Kenyan and UK faculty members were paired to work together, putting the Kenyan members’ greater contextual awareness to good advantage. Each 2.5-hour session contained between four to six separate group discussions of durations between 20 and 50 minutes. Faculty members moved between groups for each discussion to provide participants with a range of pedagogical approaches. Fifty-three participants attended the face-to-face component. Subnational travel restrictions in place due to the Covid-19 pandemic and weather-related problems prohibited the rest from attending.

### Data analysis

For each assessment tool, the raw scores were converted to percentage scores before analysis. Participants who did not complete both assessments were excluded. Improvement in score was measured as a percentage of the possible improvement, e.g., if the pre-training score was 40% and post training it was 70% then the improvement was half of the maximum potential improvement to 100% of 60%, making the improvement score 50%. Where the score decreased the change was relative to the maximum possible reduction. Relative percentage change scores were used to compare performance post-training with pre-training, by using a one sample t-test with the null hypothesis that the mean change was zero. The 5% significance level was used to determine statistical significance.

All cost data were collected in Pounds Sterling. Analysis was conducted in Microsoft EXCEL (Microsoft Corporation, Redmond, WA, USA). All costs for delivery of each version of the training were collected, these include direct participant costs as applicable (cost of travel for participants and faculty for face-to-face components, daily subsistence allowance, cost of meals during face-to-face training, cost of airtime and Kenya based faculty allowance). All UK faculty volunteered their time, so no cost was allocated to this. The cost of training equipment was not included, as the same set was used for both versions of the training. We modelled the cost for the full 5-day face-to-face approach assuming no UK faculty were required travel to support the Kenya-based faculty. We compared the mean cost per participant for both versions of the training.

### Ethical considerations

Delivery and evaluation of the AOAC using both the traditional and the blended learning approaches was approved by the Kenya Ministry of Health and all participating counties. Completing assessments before and after the training was considered part of the learning experience but participants could opt out of the assessment if they so desired after reviewing the information provided at registration. All participants were allocated unique identifying numbers and no participant identifying information was used to report the findings from this study.

## Results

### Participant satisfaction

The levels of satisfaction reported with all three elements of the blended learning were high throughout, and not significantly different from each other.

Sixty participants completed a questionnaire concerning their satisfaction regarding each facet of the course that they had attended. Of those who responded, 55 (91.7%) felt satisfied with the feedback provided as to their progress through the course and 57 (95%) were happy with the level of support provided by the education consultant.

23 (38.3%) participants reported intermittent internet connectivity problems. More than half of all participants (56.6%) reported that they were called away to attend to emergency cases during the sessions, although, despite this, 46.7% of the participants reported that they were able to concentrate on the sessions without distractions. Overall, 40 (66.7%) of participants said it was difficult to get time away from work to join the sessions. When asked if they preferred the on-line discussions to face to face training, 27 (45%) expressed a preference for on-line teaching. One participant stated, “Face to face learning is good but I find on-line discussions are able to achieve the same, maybe we just need to get used to it.” However, when those who attended the 1.5 day face-to-face practical skills training were asked at the end about their preferences, 84.9% then stated a preference for face-to-face training. This difference may have been due to the nature of the sessions, as the face-to-face sessions focusing on practical skills acquisition may have been preferable.

Several participants suggested that on-line sessions in the evenings might be easier to attend as compared to those held during normal working hours. It was apparent that many participants experienced a degree of conflict between concentrating on the sessions and the distraction of providing clinical care that would have been avoided had the participants been able to book time off to attend a course away from the workplace.

Of those responding to the survey, 58 (96.7%) felt that the content of the self-directed learning assignments was useful for their work and 57 (95%) thought that engaging with these sessions was a good use of their time. 59 (98.3%) said they had learned new things that had made them consider implementing changes to their working practices and they would like to share the contents with their work colleagues. One participant stated, “I am sorry that I did not have interest in doing the self-directed part initially, but I am now enjoying and appreciating it.” Another remarked “It was clear and precise. It was very useful to me, and I will still revisit the lectures as a reminder to myself for better practice.”

For the on-line facilitated workshops, 59 (98.3%) felt the content was relevant and useful and would recommend it to a colleague. 58 (96.7%) felt attending the sessions was a good use of their time and would be implementing changes to their practice consequently.

Of those who attended the face-to-face element of the course, 53 (100%) felt the training was relevant and useful, and that they had learned things that would change their clinical practice. All intended to increase their use of assisted vaginal delivery as a strategy to manage the second stage of labour in preference to performing a caesarean and all felt that their management of obstetric anal sphincter injuries would improve. 52 (98.1%) felt it had been a good use of their time and would recommend the training to a colleague. Across the three components of the course, 60 (100%) reported that they were either extremely or very satisfied with the overall quality of the training, and all would recommend it to other doctors. All participants agreed that the course was useful in improving the quality of care provided to patients.

Participants were asked to state the three most important things they had learned. Of those who had attended the face-to-face training, the three most commonly chosen aspects were assisted vaginal delivery techniques, newborn resuscitation and obstetric anal sphincter injury repair. Techniques for improved caesarean surgery, especially for delivering a deeply impacted fetal head, also scored highly, as did maternal resuscitation and techniques for performing venous cutdown. When asked what changes they would most like to implement, increasing the use of assisted vaginal delivery was by far the most popular, followed by consistent use of the WHO Safe Surgical Check List and a Maternal Early Obstetric Warning Scoring (MEOWS) chart using vital signs to aid prompt recognition of a deteriorating patient and track their progress in response to interventions. Several participants commented that they intended to train others in the techniques they had learned.

The participants who were unable to attend the face-to-face component reported a similar range of important lessons learned and their choices for implementation included using the MEOWS chart, use of the WHO Safe Surgical Check List and implementation of regular standards-based clinical audits in their facilities.

### Participant engagement

Sixty-five doctors engaged to some extent with the self-directed learning component of the blended training approach. Of these, 42 (64.6%) completed all 21 individual assignments within this component. A further 11 (16.9%) completed 90% or more. Four (6.2%) participants completed more than half of the assignments and 8 (12.3%) completed less than half.

We tracked the maximum time that participants spent watching the recorded lectures and videos contained within each assignment by monitoring the time taken between completing successive assignments. Whilst this did not necessarily reveal exactly how long participants spent on the respective assignments, we were able to spot those who appeared to skim through sequential assignments without adequate viewing time, or those who tried to attempt the quiz without watching the videos. Those assignments where more participants spent less than an appropriate amount of time watching included the final three assignments on the course; post-operative analgesia and anaesthetic follow up, counselling for the next birth and documentation, audit and quality of care, with 75%, 75% and 66% full engagement respectively. Other less popular assignments were safe anaesthesia (86% full engagement), safe blood transfusion (83%), managing the second stage of labour (88%) and surgical techniques and best practices for caesarean section (82%).

In total, 60 participants logged on for all or part of the five on-line discussion groups, that took place daily over five days. 54 (90%) of participants logged on for four or five days, with 38 (63.3%) present on all five occasions.

### Change in knowledge and skills

After each individual assignment participants were invited to complete a short quiz to assess their learning. The pass mark was set at 70%, and participants were provided with an explanation as to the rationale for the correct answer and offered the opportunity to repeat the test. When asked if and why they had repeated any tests, 18 (27.7%) participants said they wanted to improve their knowledge, 20 (30.8%) stated that they wanted to pass the assignment and a further 7 (10.7%) wanted to achieve a better mark. 57 (87.7%) participants had read the explanatory comments and of these, all but one found them useful.

All participants were invited to take part in a pre and post course MCQ knowledge test on-line. 61 participants completed both components. All 53 participants who attended the face-to-face component took part in Objective Structured Clinical Examinations (OSCEs) before and after the two days of training. The OSCE topics used were maternal airway management, newborn resuscitation and assisted vaginal birth. Improvement in scores occurred in both the on-line knowledge tests and OSCEs. The improvements in OSCE scores were more marked for those participants starting from a lower baseline.

Before training the mean (95% CI) MCQ score was 67% (65%-68%), whereas the mean total OSCE score was only 41% (39%-43%). For each of the OSCEs the mean (95% CI) score was similar, ranging from 39% (37%-42%) for the airways OSCE and 43% (40%-46%) for neonatal OSCE (Table 2).

**Table 2:** Comparison of knowledge and skills percentage scores post training with pre-training.

| Tool | Mean (95% CI) score |  | Relative improvement<br>(95% CI) | p-value |
| --- | --- | --- | --- | --- |
|  | Pre-training | Post-training |  |  |
| MCQ | 67 (65-68) | 84 (83-85) | 50 (47-54) | <0.001 |
| Total OSCE | 41 (39-43) | 85 (83-86) | 74 (73-76) | <0.001 |
| Assisted Vaginal Birth OSCE | 41 (38-43) | 77 (75-80) | 61 (57-65) | <0.001 |
| Neonatal resuscitation OSCE | 43 (40-46) | 91 (90-92) | 83 (80-86) | <0.001 |
| Airway management OSCE | 39 (37-42) | 86 (84-87) | 76 (73-78) | <0.001 |

There were five instances of a reduction in score after training compared with before training. Four were for the MCQ and one for the AVB OSCE, otherwise all participants OSCE scores and MCQ scores did not decrease. Figure 1 summarises the distribution of relative improvement scores.

**Fig 1.**
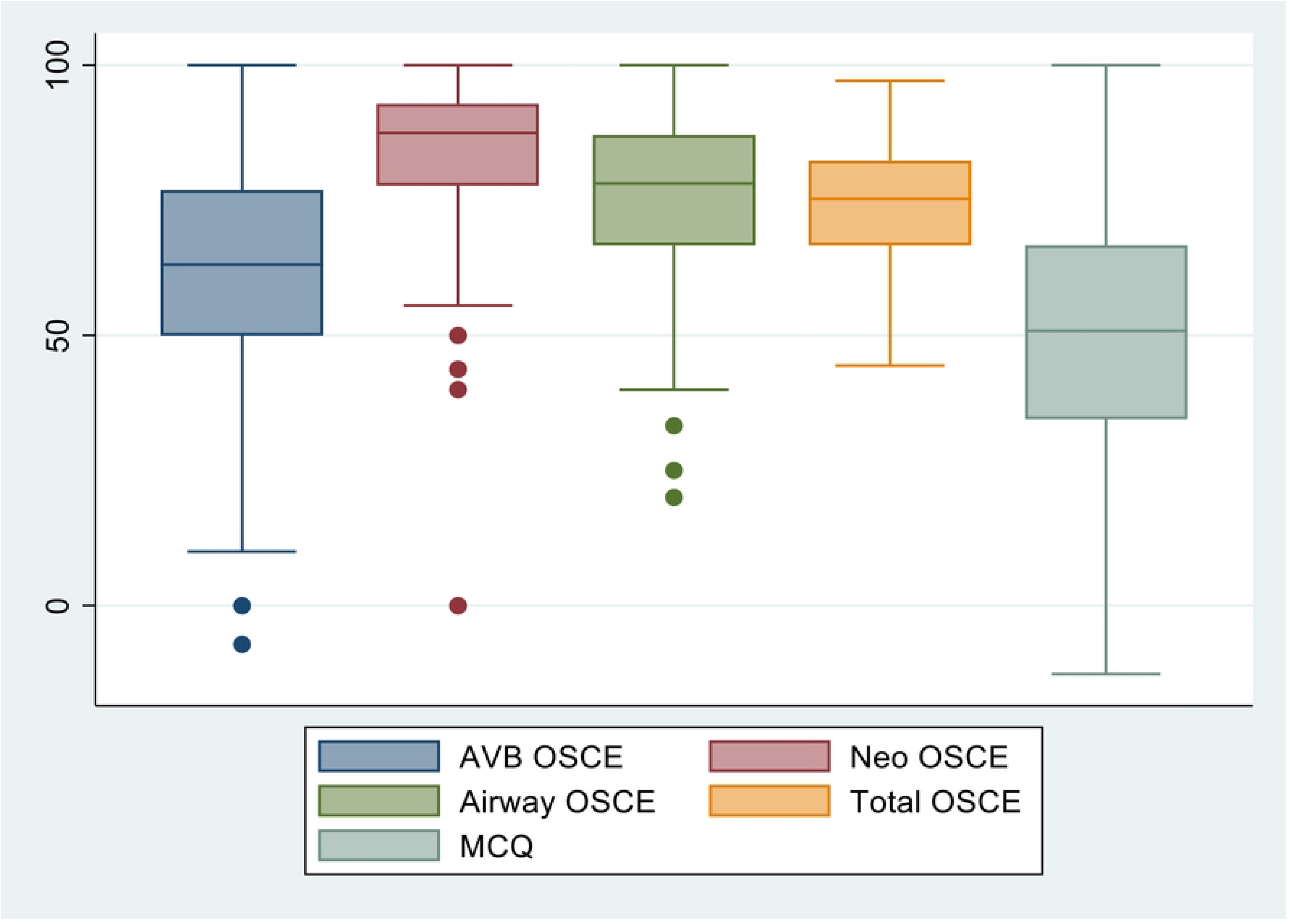
Boxplots of the distribution of relative changes in scores for each assessment tool.

After training there were statistically significant relative improvements (p<0.001 in each case) for MCQs and for each OSCE, compared with before training (Table 2). For MCQs, which had a higher percentage score before training, the mean (95% CI) relative improvement was 50% (47%-54%). For OSCEs the relative improvements were higher, with the mean (95% CI) being 61% (57%-65%) for AVB, 76% (73%-78%) for maternal airway management and 83% (80%-86%) for neonatal resuscitation.

### Comparison of cost of both approaches

The allowance for each Kenyan facilitator was in line with the Kenyan MoH guidelines, £56.10 per day for the face-to-face aspects of the training and £28.57 per day for the virtual workshop facilitation.

With regard to the practical skills training, in the blended approach 6 Kenya facilitators trained 53 participants in 3 groups, each group trained over 2 days. On the fully face-to-face course the same number of participants would have been trained in 2 groups, each trained over 5 days. The participant costs for the face-to-face component of the blending learning approach were £15,051 and the same cost for a full 5-day face-to-face course would have been £25,844. Therefore, the blended training approach costs £207 less per participant (assuming 6 facilitators and 53 participants for both courses) compared to the full 5-day face-to-face training approach.

## Discussion

We have demonstrated the feasibility of converting a five-day face-to-face advanced obstetric surgical course into a blended learning course in Kenya, reducing the face-to-face component to 1.5 days. The training was well received by most participants and both knowledge and practical skills demonstrably improved following the course. The cost saving per participant for our course was £207 (USD 250, Euro 236) after considering hotel and conference centre costs and funding for airtime. This represents a cost saving of 40% as compared to our fully face-to-face course, excluding travel expenses, which would have been incurred with either method of course delivery. Our course also resulted in a 3-day reduction of the time that staff are away from their workplace. Similar cost savings have been reported in association with another blended learning training approach (21).

There is a growing body of literature evaluating the acceptability and outcomes of blended learning in healthcare training, although direct comparisons of effectiveness are complicated by the heterogenicity of the blended learning methods employed. A systematic review by Valée et al, comparing traditional learning methods to blended learning in medical education, concluded that blended learning demonstrated consistently better effects on knowledge outcomes when compared with traditional learning in health education (22). However, of the 56 studies included in this review, only one was conducted in sub-Saharan Africa (23) and only one concerned midwifery or obstetrics (24).

The training we have described was introduced as a pragmatic solution to the challenges posed by the Covid-19 pandemic to in-service training, rather than as part of a study comparing the blended learning method of training to a fully face-to-face course. We therefore are not able to provide evidence as to whether the results we obtained from a blended learning delivery demonstrated greater, the same or lesser improvement in knowledge and skills acquisition. Nevertheless, we have been able to demonstrate a highly significant relative improvement in both knowledge and skills of participants between before and after tests, with improvements comparable to a before and after study of knowledge and skills from a multi-country study of face-to-face emergency obstetric skills training (14).

Our choice of on-line platform for the self-directed component was made taking into consideration the need for sustainability. Although some bespoke platforms would have been easier to work with, we prioritised choosing a platform that was available at no cost because we wanted it to remain freely available in the future to participants. If courses such as this are to be available to health workers in low resource settings maintaining access at zero or low cost is important. Following the pilot of the blended learning delivery approach, we have now hosted the self-directed learning component on the World Continuing Education Alliance (WCEA) platform. Health worker professional and regulatory bodies in most of sub-Saharan Africa have validated courses on this platform for continuous professional development (CPD). All courses on the WCEA platform are available free of charge to health workers from low-and-middle income countries.

One disadvantage of providing internet-based training remains the cost of purchasing air time. Although the internet is widely available, many health workers in low resource settings do not have contracts with unlimited internet availability and they may struggle to fund air time for training unless provided with financial support. We did fund air time for this course, but this would inevitably affect sustainability if funding was not provided. However, in several countries in sub-Saharan Africa CPD is increasingly becoming mandatory for health workers linked to practice license renewal. Policy makers need to make allowance for covering training costs as part of an overall strategy to ensure competent skilled birth personnel, a key strategy to achieving the SDG3 maternal mortality reduction target.

Aware of the difficulties that participants might encounter using an unfamiliar platform, in mitigation we provided them with a document and video explaining platform navigation step by step. This worked well for most of the participants, despite their lack of familiarity with the platform.

Tracking the extent to which participants engaged with the on-line components was key to our evaluation of this course. Some participants stated in feedback that they tended to avoid certain topics because they had judged that they already knew enough about these topics. Other participants explained that they had watched the videos throughout but had exploited the facility on the platform to watch at a faster than normal speed. The ability to pick and choose which aspects of the course to focus on could represent a good use of participant’s time, although this does rely on the accuracy of their judgement as to the adequacy of their knowledge.

We had explained to participants that the self-directed component would take approximately 15 hours to complete, and assignments were released to the participants two weeks prior to the on-line discussion group component. Nevertheless, some participants felt that they did not have sufficient time to complete all the assignments, commenting that they struggled to complete the lectures in between reporting daily for work and having very short times off duty due to staff shortages. 10 (15.4%) of the participants reported that the self-directed assignments took longer to complete than they had expected. Earlier release of the self-directed component to participants may reduce time pressures in future.

It was clear that a significant number of participants found it difficult to book time off from their work commitments to enable them to attend the on-line discussion groups. Learning in the discussion groups may have been compromised by participants becoming distracted by on-going clinical issues in the workplace. Although we ensured that only half of the doctors attending the discussion groups from any facility were booked for any given session, nevertheless, due to staff shortages it proved impossible for some doctors to give the groups their undivided attention. The 2.5-hour duration of the sessions may have been a factor, as booking this time off work may have been more challenging when compared to booking an entire day or half day. This may have encouraged doctors to hope for the best regarding clinical workload and try to manage clinical commitments and attend the sessions simultaneously. A potential solution would be to run these sessions in the evening, depending upon acceptability to both participants and faculty. However, concerns have been expressed regarding the potential for on-line learning to invade participants’ everyday lives suggesting evening sessions may not be suitable for all participants (25).

Running the discussion groups on-line enabled UK and Kenyan faculty members to work together without the need for either international or national travel. This enabled international collaboration whilst removing the requirement for travel, providing cost savings and environmental benefits. Faculty members from both settings enjoyed working in collaboration and learned from each other.

Although, in the context of the Coronavirus pandemic, converting 100% of the training to internet based would have been preferable, it was considered that it would have been very challenging for participants to acquire the necessary clinical skills in the absence of the opportunity for hands on practice that the face-to-face element provided. Some skills improvement may occur from watching video demonstrations, but supervised hands-on practice using high fidelity models has a track record of success (14) and this was replicated here as evidenced by the improvement in OSCE scores.

Of concern were the low pre-course OSCE scores for basic clinical skills across all cadres. The medical officers’ and interns’ scores were low across all OSCEs. Medical officers form the backbone of service provision in many facilities, especially at sub-county level, where they provide obstetric care, usually without senior supervision. From our initial assessment, none of the medical officers were able to demonstrate basic clinical skills sufficient to safely manage a maternal airway and provide ventilation using an Ambu® bag and mask, and only one was able to demonstrate competency in newborn resuscitation. Similarly, only one medical officer had appropriate AVD skills. This may explain the reticence to conduct assisted vaginal birth seen in some Kenyan health facilities (26). Although doctors in Kenya are expected to provide evidence of CPD, these findings would suggest that a more prescriptive approach to the nature of CPD activities would be of benefit, including regular clinical skills training for doctors at all levels, as is the case in some other countries where mandatory training includes regular updates on basic resuscitation skills. This training has demonstrated a significant uplift is skills after a short intervention. Although we have not been able yet to demonstrate sustained improvement, previous evidence suggests that skills are retained for at least a year (15). Participants themselves clearly recognised the need to improve the provision of assisted vaginal birth in their units, and to practice and teach improved resuscitation skills for both mothers and babies, as judged by their stated intentions on finishing the training.

### Limitations

We acknowledge that it is not possible to report the exact degree of engagement with the on-line components of this training. However, even when sitting in a face-to-face lecture, it is possible for the participant’s mind to wander, hence attention cannot be directly measured in either situation. This study did not set out to measure comparative effectiveness between traditional face-to-face and blended learning teaching methods but rather the acceptability and feasibility of blended learning in the context described. We acknowledge that it will be important to compare the impact of both versions of the training on maternal and newborn health outcomes.

Across the cadres, the majority of those recruited were medical officers, with smaller numbers of the other cadres and the results for these cadres may not be generalisable. While the content of the training was more suitable for medical doctors, midwives and anaesthetists are involved in peri-soperative and postnatal care, and a multidisciplinary training is likely to facilitate change in practice and behaviour.

## Conclusion

We have demonstrated that a blended learning approach to clinical training in a low resource setting is feasible and sustainable, resulting in a reduction in time taken for face-to-face training, with this component being reserved for practical skills acquisition. We recommend that consideration is given to the development of a more structured approach to continued professional development for doctors providing obstetric services, to include mandatory clinical skills updates on a regular basis.

## Data Availability

All data relevant to the study are included in the article.

